# COVID-19 and p*er capita* green tea consumption: update

**DOI:** 10.1101/2022.06.06.22276060

**Authors:** Maksim Storozhuk

**Author notes:** Corresponding author Dr. M. Storozhuk, Bogomoletz Institute of Physiology, National Academy of Science of Ukraine, 4 Bogomoletz Street, Kyiv 01024, Ukraine.

## Abstract

**Purpose:** In spite of the development of numerous vaccines for the prevention of COVID-19 and approvement of several drugs for its treatment, there is still a great need in effective and inexpensive therapy of this disease. Pharmacological evidence suggesting the therapeutic potential of green tea catechins in amelioration/treatment of COVID19 is growing rapidly, however, there are only a few epidemiological studies addressing this possibility. The aim of this study was to provide update regarding ecological study assessing this issue as of January 2021.

**Methods:** The methodological approach used in this report is similar to that described previously. Briefly, information about COVID-19 morbidity (defined as a total number of cases per million population) and mortality (defined as a total number of deaths per million population) for a specific date was directly obtained from Worldometers info. Coronavirus. Analysis was restricted to 134 countries or territories with at least 3 million population. Twenty-one of these countries/territories, with estimated *per/capita* green tea consumption above 150 g (annually), were considered as a group with the high consumption. Countries/territories with the estimated *per/capita* green tea consumption below 150 g (N=82) were considered as the group with low the consumption.

**Results:** *P*ronounced differences in COVID-19 morbidity and mortality between groups of countries with high and low green tea consumption were found as of February 20, 2022. These differences were still observed in a subset of countries with HDI above 0.55. Moreover, in this restricted subset of countries, weak but statistically significant correlations between COVID-19 morbidity (or mortality) and per/capita green tea consumption were observed in a multiple regression model accounting for: population density, percentage of population aged above 65, and percentage of urban population.

**Conclusion:** The obtained results provide additional, though indirect, support of the idea that green tea catechins can be useful for treatment/amelioration of COVID-19. These results are in line with emerging evidence from other studies, including pharmacological. Nevertheless, further research is necessary to directly validate or reject this idea.

## Introduction

In spite of the development of numerous vaccines for prevention of COVID-19 and approvement of several drugs for its treatment, there is still a great need for effective and inexpensive therapy of this disease. In this regard therapeutic potential of green tea catechins looks promising. There is mounting evidence suggesting the therapeutic potential of green tea catechins in prevention/treatment of COVID19. In particular, in addition to numerous docking studies (for instance, [1-7]) there is growing number of studies showing direct antiviral activity of at least one catechin epigallocatechin-3-gallate (EGCG) against SARS-CoV-2 in *in vitro* experiments ([8-17] see also [18-20] for review). Moreover, a recent work [21] reports that EGCG from green tea effectively blocks infection of SARS-CoV-2 and *new variants*. The latter is in line with the observation [22] that neutralizing activity of concentrated green tea extract is independent of the strain of SARS-CoV-2 (Wuhan strain, beta- or delta-variants).

Besides, green tea constituent are likely to be beneficial in relation to factors associated with higher COVID-19 mortality such as cholesterol levels [23], obesity [24], [25], diabetes [26], uncontrolled immune activation, [27], and cardiovascular disease [28] Finally, green tea catechins potentiate the adaptive immunity [12] and can act as ionophores for zinc ions, while the latter are considered as potentially beneficial in relation to COVID-19 [29].

Still, it appears that there are only a few epidemiological studies assessing therapeutic potential of green tea catechins: (i) observational study [30] reporting that people who consumed ≥4 cups/day of green tea had a lower, albeit statistically not significant, odds of SARS-CoV-2 infection; (ii) ecological studies [31], [32] reporting lower COVID-19 morbidity and mortality in countries with higher *per capita* green tea consumption.

Additionally, although differences in COVID-19 morbidity and mortality between groups of countries with higher and lower *per capita* green tea consumption were statistically significant in the abovementioned ecological studies, data analyzed in these studies reflected the epidemiological situation observed in January 2021 and before. The aim of this report was to examine whether differences in COVID-19 morbidity and mortality between countries with higher and lower *per capita* green tea consumption are still present in the year 2022.

## Methods

### Ethics approval and consent to participate

Not applicable (ethical approval was not deemed necessary as this was an analysis of publicly available data).

### Data regarding COVID-19 morbidity and mortality

All data were obtained from open sources. Information on the total number of cases and total number of deaths was obtained directly from ‘Worldometers info. Coronavirus’ [33]. The information on ‘Worldometer’ is based on official daily reports and considered as a reliable (for instance, [34-35]).

The methodological approach used in this report is similar to that described previously [31-32]. Briefly, information about COVID-19 morbidity (defined as a total number of cases per million population) and mortality (defined as a total number of deaths per million population) for a specific date was directly obtained from ‘Worldometers info. Coronavirus’ [33]. Analysis was restricted to 134 countries or territories (according UN classification) with at least 3 million population. Twenty-one of these countries/territories, with estimated *per/capita* green tea consumption above 150 g (annually), were considered as a group with the high consumption. Countries/territories with estimated *per/capita* green tea consumption below 150 g were considered as a group with the low consumption (see [31-32] for details). Considering that the variables of COVID-19 morbidity and COVID-19 mortality do not have a normal distribution [34], non–parametric statistic (Wilcoxon (Mann-Whitney U Test) for Unpaired Data) was used for comparisons.

In multiple linear regression analysis, the following factors (beside green tea consumption) were included: population density [35], percentage of population aged above 65 [36], percentage of urban population [37]. In a complementary analysis an additional variable, namely Human Developmental Index (HDI) based on access to health and education services and income [38] was added to the model. ‘KyPlot’ software was employed for statistical assessments.

## Results

Pronounced and statistically significant differences in COVID-19 morbidity and mortality between groups of countries/territories with higher and lower green tea consumption were found as of February 20, 2022 (**Table 1**). Thus, it appears that previously reported differences in COVID-19 morbidity and mortality, reflecting epidemiological situation in January 2021 [32] and before [31], are consistent over a prolonged time. This consistency may be relevant to the reported efficacy of green tea catechins against *several* variants of SARS-CoV-2, which was observed in *in vitro* studies [21-22].

**Table 1.**
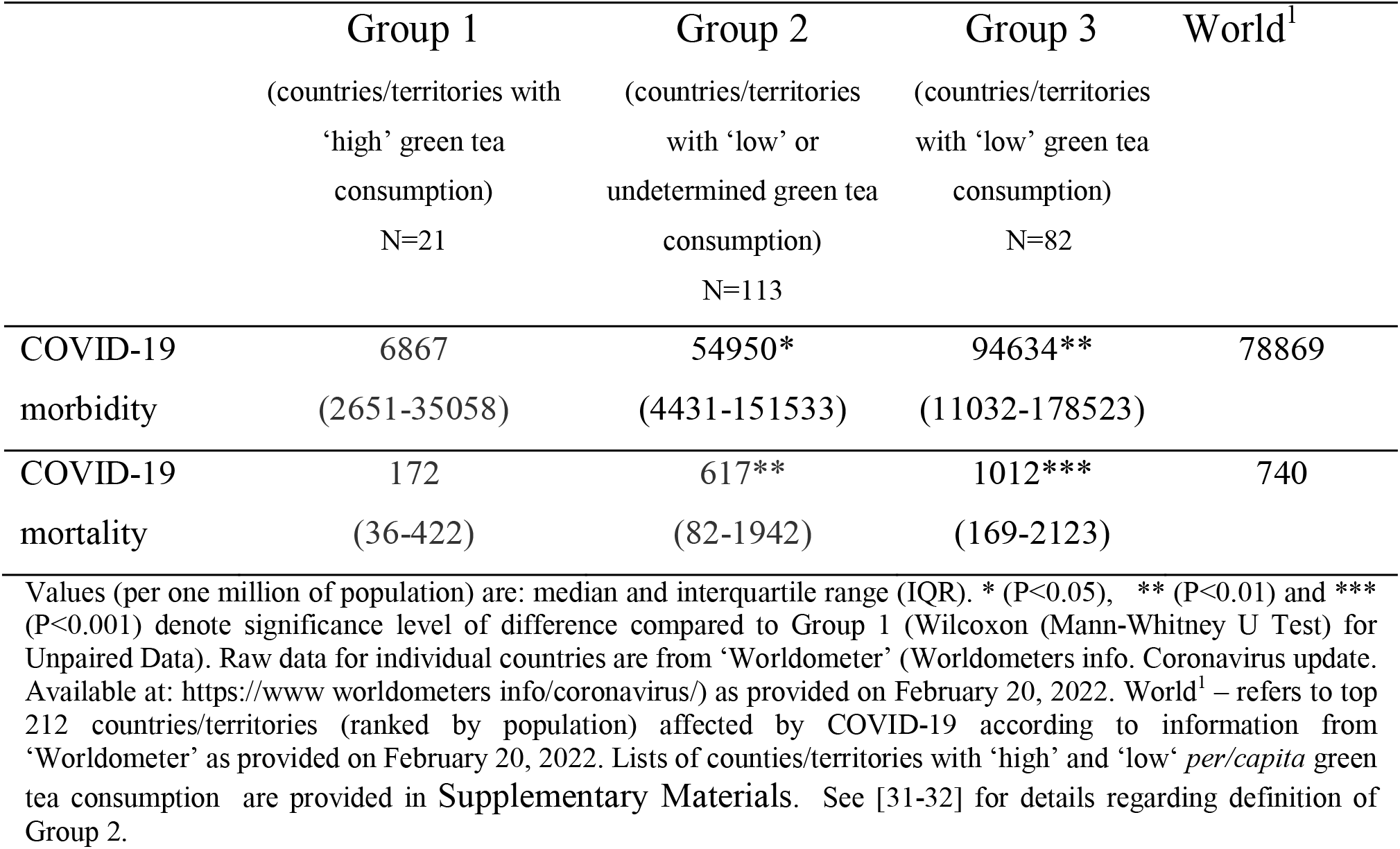
Lower COVID-19 morbidity and mortality in countries with higher *per capita* green tea consumption.

One of the factors strongly associated with COVID-19 morbidity and mortality is HDI [39]. Indeed, for several reasons (including underestimation of COVID-19 morbidity and mortality due to lower COVID-19 testing capabilities) COVID-19 morbidity and mortality appear to be lower in countries with lower HDI. This is especially apparent in countries with low HDI (countries with HDI below 0.55, as defined by UN) such as Burundi, Burkina Faso, Madagascar, Niger. Thus, it may be argued that lower COVID-19 morbidity and mortality in the group of countries with higher per capita green tea consumption is biased due to overrepresentation of countries with low HDI in this group. However, if countries with HDI below 0.55 were excluded from the analysis, there were still large and statistically significant differences between groups with high and low green tea consumption (**Table S1)**.

Moreover, in this restricted subset of countries, weak but statistically significant correlations between COVID-19 morbidity (or mortality) and per/capita green tea consumption were observed in a multiple regression model accounting for several factors, reported previously [39] as important confounders (population density, percentage of population aged above 65, percentage of urban population). These results are summarized in the **Table S2 (A** and **B)**.

Although vaccination alone is not sufficient to contain the outbreak [40], potentially, differences in vaccination rates against COVID-19 in distinct countries can be another source of systematic bias. On the other hand, the period of time analyzed here (since the beginning of epidemic in December 2019 to February 2022) includes both pre- and post vaccination parts since the earliest mass vaccination was initiated in the second half of December 2020. Besides, a reasonable assumption is that vaccination rates (during post vaccination period) are associated with HDI. Therefore, in part, this concern may be addressed by adjusting for HDI. **Table S3** shows that correlations between COVID-19 morbidity (or mortality) and per/capita green tea consumption are still statistically significant in multiple regression model accounting *also* for HDI.

Interestingly, the transformation of the variable per capita green tea consumption into the common logarithm Log(10), notably increased the strength and statistical significance of the correlations between this variable and COVID-19 morbidity or mortality (see the values in **Table S4** compared to those in **Table S3)**. Apart from other reasons not discussed here, these increases might be relevant to the logarithmic dependence between dose and effect, usually observed in pharmacological studies.

## Discussion

Pronounced differences in COVID-19 morbidity and mortality between groups of countries/territories with higher and lower green tea consumption were found as of February 20, 2022. These results extend previous observations, reflecting the epidemiological situation in January 2021 [32] and before (September and November 2020 [31]). This consistency over prolonged period of time may be an additional argument supporting the therapeutic potential of green tea catechins in amelioration/treatment of COVID19.

Although ecological studies, taken alone, could not confirm a causal relation, still these studies are considered as useful and widely used in the field, in particular, in relation to COVD-19 (e.g., [34], [39]. The current report, as well as ecological studies in general, does have limitations. In particular, there are many factors that can differentially affect COVID-19 morbidity and mortality in distinct countries (e.g. the percentage of older population; HDI [39], administrative strategies to prevent COVID-19 transmission; preventive strategies related to *other* diseases [39], [34], [41]; condition-specific mortality risks; particularities of treatment and vitamin D supplementation [42]). Additionally, particularities of the tea predominantly consumed in a specific country may also be of importance in this respect, because tea processing conditions can affect the content of potential antiviral compounds [43]. On the other hand, since numerous (more than one hundred) countries from all over the world were considered, it does not seem likely that these factors can *systematically* and *strongly* bias the results presented here. To some extent, this point is supported by similarities of the results obtained without restriction in respect of HDI (**Table 1**), and when countries with low HDI were excluded (**Table S1**). Additionally, this point is supported by the results of linear regression analysis accounting for several factors, reported previously as important confounders in relation to COVID-19 morbidity and mortality. Indeed, statistically significant correlations between COVID-19 morbidity and mortality and per capita green tea consumption were observed in the linear regression analysis accounting for the following: population density, percentage of population aged above 65, percentage of urban population, and HDI (**Table S3**).

In any case, although these results do not *necessarily* indicate the causal links between higher green tea consumption and lower COVID-19 morbidity/mortality they do support these potential links. And this is in agreement with emerging pharmacological evidence ([1], [8], [9], [18], [21]) and the trend observed in the above-mentioned observational study [30].

Currently, at least one placebo controlled study has been registered to access potential of EGCG-containing formulation (taken internally) in prevention of COVID-19 [44], but the results have not yet been reported. Interestingly, additional approach regarding green tea catechins and COVID-19 is also emerging [22]. This approach [22] focuses on the *local application* of green tea extract as a throat spray. It was demonstrated that the green tea extract formulation has strong neutralizing activity on SARS-CoV-2 independent of the strain (Wuhan strain, beta- or delta-variants) in VeroE6 cell culture model. Additionally qualitative and quantitative tannin profile present on the oral mucosa after spray application has been investigated [22]. In relation to the ecological data discussed here, it seems to be of interest to study whether usual green tea consumption also substantially enhances the level of green tea constituents in oral mucosa. Actually, if links between higher green tea consumption and lower COVID-19 morbidity/mortality are indeed causal, it cannot be excluded that this is (in part) due to *local* action of green tea catechins in mouth/throat. If this is the case, another aspect of this issue arises. Should lower efficacy of green tea catechins formulations be expected in clinical trials, in which these formulations are taken *internally as capsules* (for instance, as compared to observational studies with a design similar to [30])? How to address this potential issue in placebo-controlled studies?

To conclude, the obtained results provide additional though indirect support of the idea that green tea catechins can be useful for treatment/amelioration of COVID-19. These results are in line with the rapidly growing evidence obtained in pharmacological *in vitro* studies. Nevertheless, further research is necessary to directly validate or reject this idea.

## Data Availability

All data produced in the present work are contained in the manuscript

## Declarations

### Ethical Approval and Consent to participate

Not applicable.

### Consent for publication

Not applicable

### Availability of supporting data

Data (mostly) available within the manuscript and supplementary materials. Additional data are available on request from the author.

## Acknowledgments

I would like express my gratitude to personnel of the open resources for creating excellent opportunities for research.

## Supplementary Materials

*List of countries/territories with ‘high’ per/capita green tea consumption (ranked by population)*. China, Indonesia, Japan, Vietnam, Algeria, Afghanistan, Uzbekistan, Morocco, Niger, Taiwan, Burkina Faso, Mali, Senegal, Guinea, Tunisia, UAE, Hong Kong, Libya, Denmark, Mauritania, Mongolia.

*List of countries/territories with ‘low’ per/capita green tea consumption (ranked by population)*.

USA, Pakistan, Brazil, Nigeria, Russia, Mexico, Ethiopia, Philippines, Egypt, Turkey, Iran, Germany, Thailand, UK, France, Italy, Colombia, Spain, Sudan, Ukraine, Canada, Poland, Saudi Arabia, Angola, Peru, Malaysia, Ghana, Madagascar, Cameroon, Ivory Coast, Australia, Malawi, Chile, Zambia, Kazakhstan, Romania, Syria, Ecuador, Netherlands, Cambodia, Zimbabwe, Rwanda, Benin, Burundi, Bolivia, Belgium, Dominican Republic, Czechia, Greece, Azerbaijan, Sweden, Honduras, Portugal, Hungary, Belarus, Israel, Austria, Switzerland, Serbia, Togo, Sierra Leone, Laos, Paraguay, Bulgaria, Lebanon, Nicaragua, Kyrgyzstan, El Salvador, Singapore, Finland, Norway, Slovakia, Oman, Costa Rica, Ireland, New Zealand, CAR, Panama, Kuwait, Croatia, Uruguay, Bosnia and Herzegovina.

See [32] for more details. These lists refer to countries with HDI above and below 0.55.

**Table S1.**
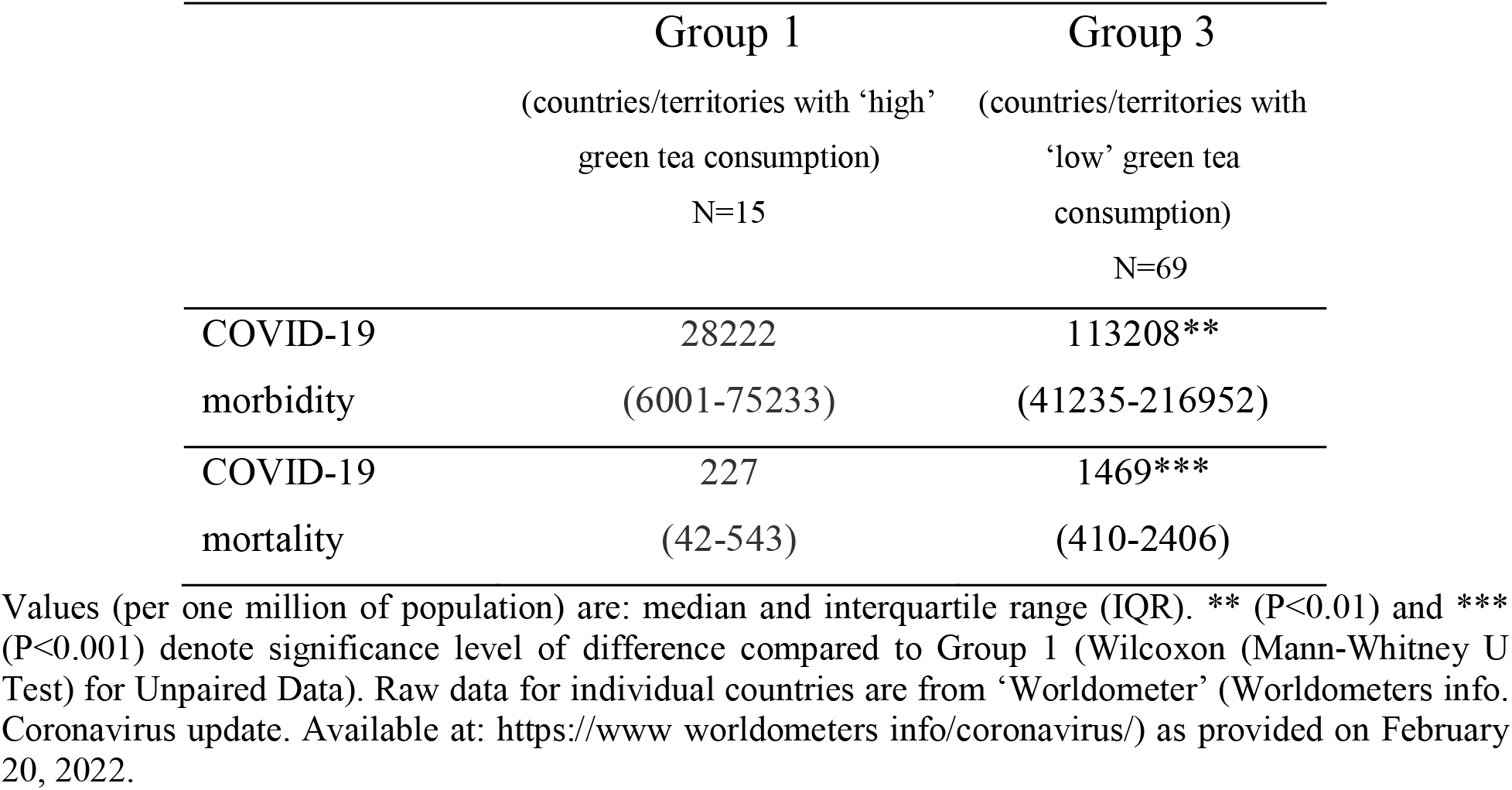
COVID-19 morbidity and mortality in relation to per/capita green tea consumption in subset of countries with HDI above 0.55

**Table S2.**
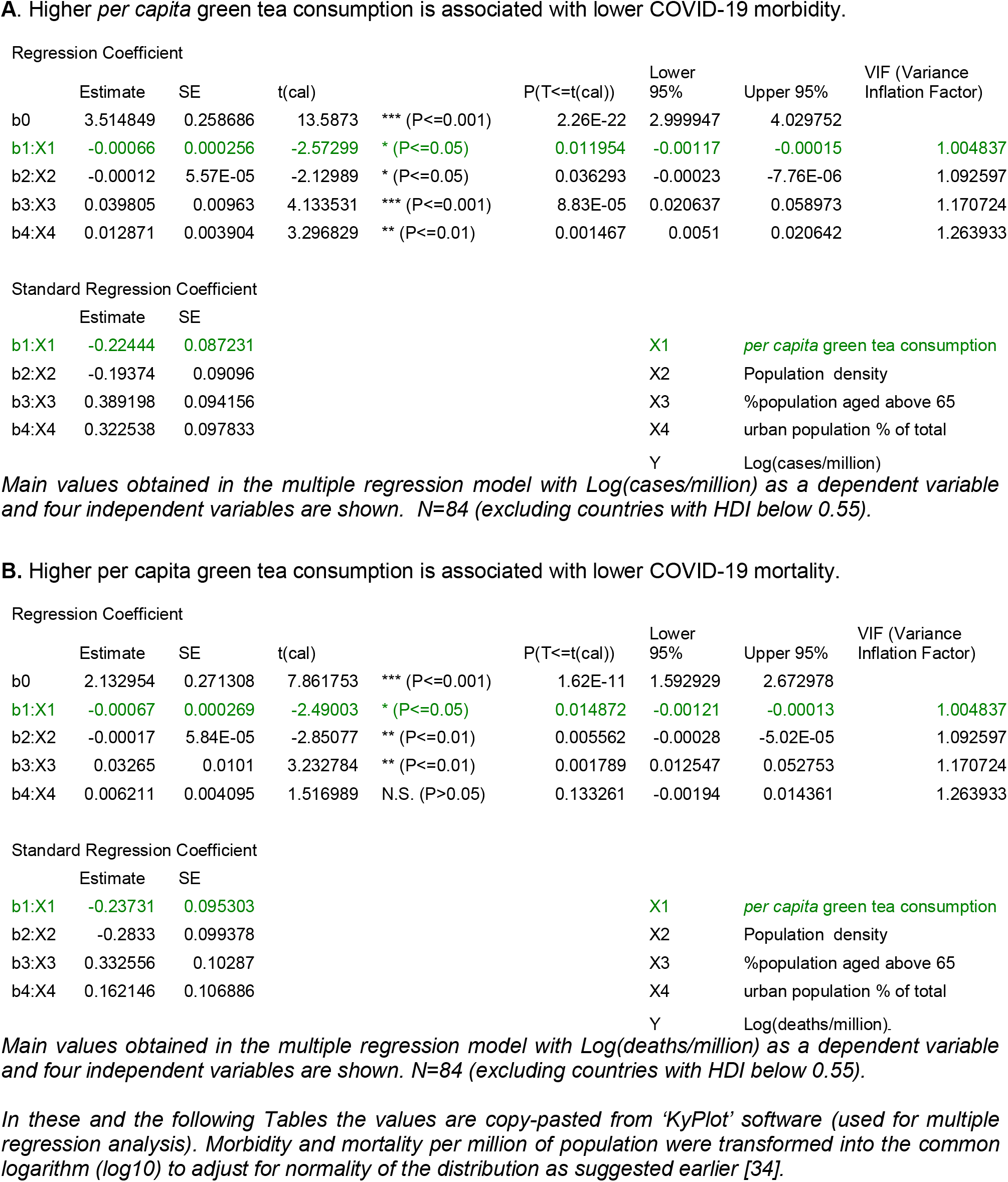
A. Higher *per capita* green tea consumption is associated with lower COVID-19 morbidity.

| Regression Coefficient |  |  |  |  |  |  |  |  |
| --- | --- | --- | --- | --- | --- | --- | --- | --- |
|  | Estimate | SE | t(cal) |  | P(T<=t(cal)) | Lower 95% | Upper 95% | VIF (Variance Inflation Factor) |
| b0 | 3.514849 | 0.258686 | 13.5873 | *** (P<=0.001) | 2.26E-22 | 2.999947 | 4.029752 |  |
| b1:X1 | -0.00066 | 0.000256 | -2.57299 | * (P<=0.05) | 0.011954 | -0.00117 | -0.00015 | 1.004837 |
| b2:X2 | -0.00012 | 5.57E-05 | -2.12989 | * (P<=0.05) | 0.036293 | -0.00023 | -7.76E-06 | 1.092597 |
| b3:X3 | 0.039805 | 0.00963 | 4.133531 | *** (P<=0.001) | 8.83E-05 | 0.020637 | 0.058973 | 1.170724 |
| b4:X4 | 0.012871 | 0.003904 | 3.296829 | ** (P<=0.01) | 0.001467 | 0.0051 | 0.020642 | 1.263933 |

| Standard Regression Coefficient |  |  |  |  |  |  |
| --- | --- | --- | --- | --- | --- | --- |
|  | Estimate | SE |  |  |  |  |
| b1:X1 | -0.22444 | 0.087231 |  |  | X1 | *per capita green tea consumption* |
| b2:X2 | -0.19374 | 0.09096 |  |  | X2 | Population density |
| b3:X3 | 0.389198 | 0.094156 |  |  | X3 | %population aged above 65 |
| b4:X4 | 0.322538 | 0.097833 |  |  | X4 | urban population % of total |
|  |  |  |  |  | Y | Log(cases/million) |

| Regression Coefficient |  |  |  |  |  |  |  |  |
| --- | --- | --- | --- | --- | --- | --- | --- | --- |
|  | Estimate | SE | t(cal) |  | P(T<=t(cal)) | Lower 95% | Upper 95% | VIF (Variance Inflation Factor) |
| b0 | 2.132954 | 0.271308 | 7.861753 | *** (P<=0.001) | 1.62E-11 | 1.592929 | 2.672978 |  |
| b1:X1 | -0.00067 | 0.000269 | -2.49003 | * (P<=0.05) | 0.014872 | -0.00121 | -0.00013 | 1.004837 |
| b2:X2 | -0.00017 | 5.84E-05 | -2.85077 | ** (P<=0.01) | 0.005562 | -0.00028 | -5.02E-05 | 1.092597 |
| b3:X3 | 0.03265 | 0.0101 | 3.232784 | ** (P<=0.01) | 0.001789 | 0.012547 | 0.052753 | 1.170724 |
| b4:X4 | 0.006211 | 0.004095 | 1.516989 | N.S. (P>0.05) | 0.133261 | -0.00194 | 0.014361 | 1.263933 |

| Standard Regression Coefficient |  |  |  |  |  |  |
| --- | --- | --- | --- | --- | --- | --- |
|  | Estimate | SE |  |  |  |  |
| b1:X1 | -0.23731 | 0.095303 |  |  | X1 | *per capita green tea consumption* |
| b2:X2 | -0.2833 | 0.099378 |  |  | X2 | Population density |
| b3:X3 | 0.332556 | 0.10287 |  |  | X3 | %population aged above 65 |
| b4:X4 | 0.162146 | 0.106886 |  |  | X4 | urban population % of total |
|  |  |  |  |  | Y | Log(deaths/million). |

*Main values obtained in the multiple regression model with Log(deaths/million) as a dependent variable and four independent variables are shown. N=84 (excluding countries with HDI below 0.55).*
*In these and the following Tables the values are copy-pasted from 'KyPlot' software (used for multiple regression analysis). Morbidity and mortality per million of population were transformed into the common logarithm (log10) to adjust for normality of the distribution as suggested earlier [34].*

**Table S3.**
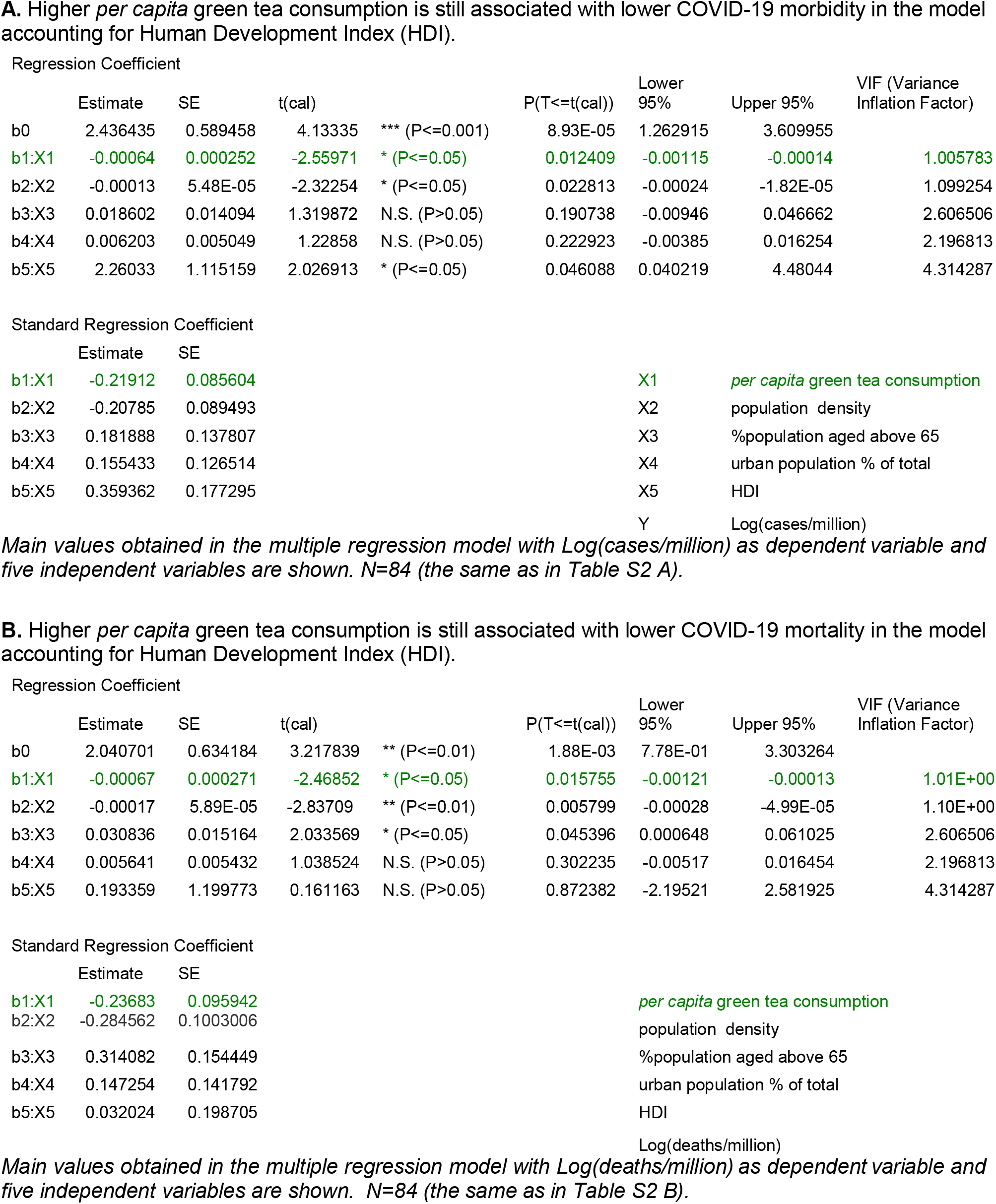

| Regression Coefficient |  |  |  |  |  |  |  |  |
| --- | --- | --- | --- | --- | --- | --- | --- | --- |
|  | Estimate | SE | t(cal) |  | P(T<=t(cal)) | Lower 95% | Upper 95% | VIF (Variance Inflation Factor) |
| b0 | 2.436435 | 0.589458 | 4.13335 | *** (P<=0.001) | 8.93E-05 | 1.262915 | 3.609955 |  |
| b1:X1 | -0.00064 | 0.000252 | -2.55971 | * (P<=0.05) | 0.012409 | -0.00115 | -0.00014 | 1.005783 |
| b2:X2 | -0.00013 | 5.48E-05 | -2.32254 | * (P<=0.05) | 0.022813 | -0.00024 | -1.82E-05 | 1.099254 |
| b3:X3 | 0.018602 | 0.014094 | 1.319872 | N.S. (P>0.05) | 0.190738 | -0.00946 | 0.046662 | 2.606506 |
| b4:X4 | 0.006203 | 0.005049 | 1.22858 | N.S. (P>0.05) | 0.222923 | -0.00385 | 0.016254 | 2.196813 |
| b5:X5 | 2.26033 | 1.115159 | 2.026913 | * (P<=0.05) | 0.046088 | 0.040219 | 4.48044 | 4.314287 |
| Standard Regression Coefficient |  |  |  |  |  |  |  |  |
|  | Estimate | SE |  |  |  |  |  |  |
| b1:X1 | -0.21912 | 0.085604 |  |  |  | X1 | *per capita green tea consumption* |  |
| b2:X2 | -0.20785 | 0.089493 |  |  |  | X2 | population density |  |
| b3:X3 | 0.181888 | 0.137807 |  |  |  | X3 | %population aged above 65 |  |
| b4:X4 | 0.155433 | 0.126514 |  |  |  | X4 | urban population % of total |  |
| b5:X5 | 0.359362 | 0.177295 |  |  |  | X5 | HDI |  |
|  |  |  |  |  |  | Y | Log(cases/million) |  |

| Regression Coefficient |  |  |  |  |  |  |  |  |
| --- | --- | --- | --- | --- | --- | --- | --- | --- |
|  | Estimate | SE | t(cal) |  | P(T<=t(cal)) | Lower 95% | Upper 95% | VIF (Variance Inflation Factor) |
| b0 | 2.040701 | 0.634184 | 3.217839 | ** (P<=0.01) | 1.88E-03 | 7.78E-01 | 3.303264 |  |
| b1:X1 | -0.00067 | 0.000271 | -2.46852 | * (P<=0.05) | 0.015755 | -0.00121 | -0.00013 | 1.01E+00 |
| b2:X2 | -0.00017 | 5.89E-05 | -2.83709 | ** (P<=0.01) | 0.005799 | -0.00028 | -4.99E-05 | 1.10E+00 |
| b3:X3 | 0.030836 | 0.015164 | 2.033569 | * (P<=0.05) | 0.045396 | 0.000648 | 0.061025 | 2.606506 |
| b4:X4 | 0.005641 | 0.005432 | 1.038524 | N.S. (P>0.05) | 0.302235 | -0.00517 | 0.016454 | 2.196813 |
| b5:X5 | 0.193359 | 1.199773 | 0.161163 | N.S. (P>0.05) | 0.872382 | -2.19521 | 2.581925 | 4.314287 |
| Standard Regression Coefficient |  |  |  |  |  |  |  |  |
|  | Estimate | SE |  |  |  |  |  |  |
| b1:X1 | -0.23683 | 0.095942 |  |  |  |  | *per capita green tea consumption* |  |
| b2:X2 | -0.284562 | 0.1003006 |  |  |  |  | population density |  |
| b3:X3 | 0.314082 | 0.154449 |  |  |  |  | %population aged above 65 |  |
| b4:X4 | 0.147254 | 0.141792 |  |  |  |  | urban population % of total |  |
| b5:X5 | 0.032024 | 0.198705 |  |  |  |  | HDI |  |
|  |  |  |  |  |  |  | Log(deaths/million) |  |

**Table S4.**
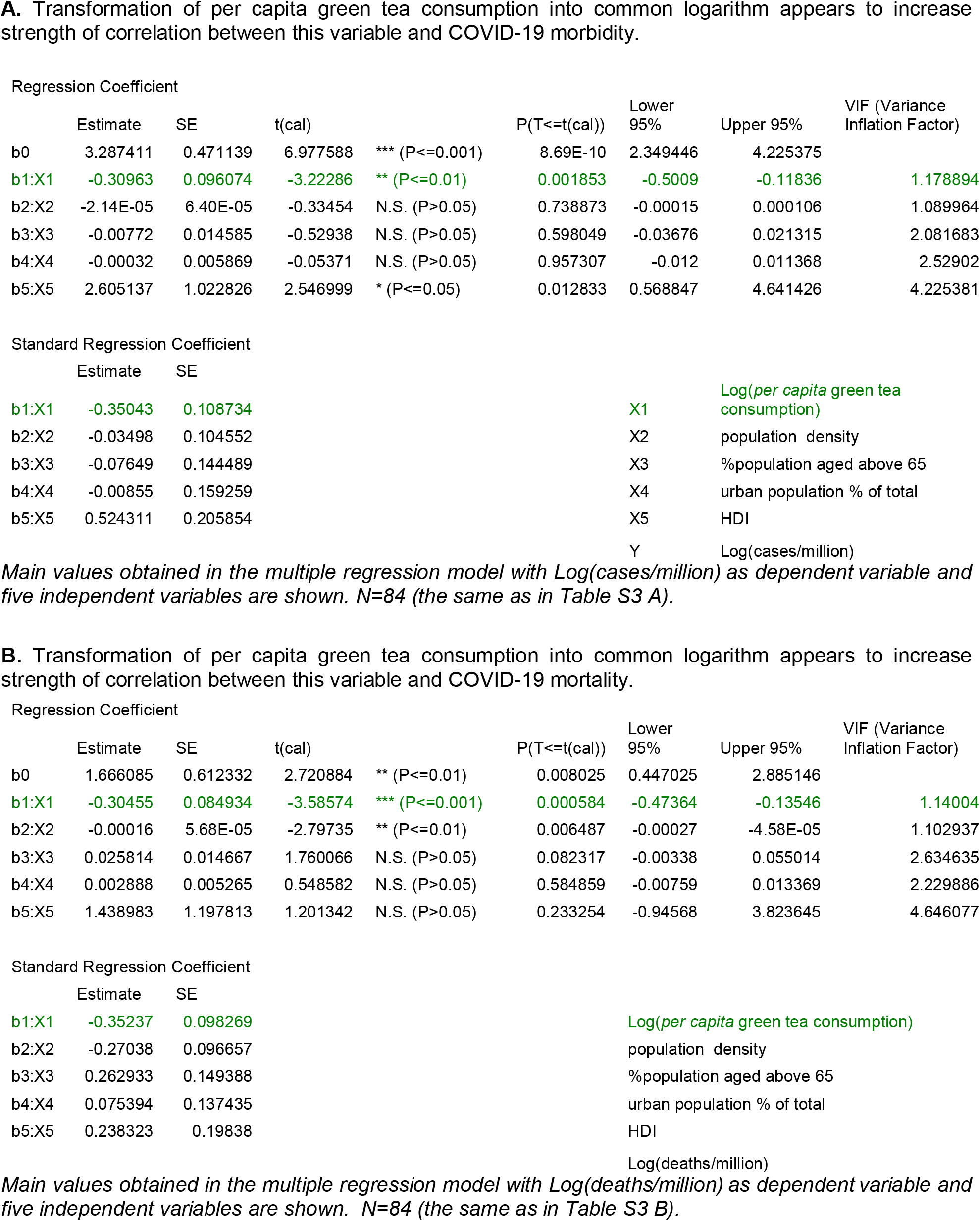

## Reference List

[1] Wu C, Liu Y, Yang Y et al. Analysis of therapeutic targets for SARS-CoV-2 and discovery of potential drugs by computational methods. Acta Pharm Sin B 2020. https://doi.org/10.1016/j.apsb.2020.02.008

[2] Sagaama A, Brandan SA, Ben IT, Issaoui N. Searching potential antiviral candidates for the treatment of the 2019 novel coronavirus based on DFT calculations and molecular docking. Heliyon 2020;6:e04640. https://doi.org/10.1016/j.heliyon.2020.e04640

[3] Allam L, Ghrifi F, Mohammed H et al. Targeting the GRP78-Dependant SARS-CoV-2 Cell Entry by Peptides and Small Molecules. Bioinform Biol Insights 2020;14:1177932220965505. https://doi.org/10.1177/1177932220965505

[4] Mhatre S, Gurav N, Shah M, Patravale V. Entry-inhibitory role of catechins against SARS-CoV-2 and its UK variant. Comput Biol Med 2021;135:104560. https://doi.org/10.1016/j.compbiomed.2021.104560

[5] Soni U, Singh P, Gupta OP et al. Lichen planus drugs re-purposing as potential anti COVID-19 therapeutics through molecular docking and molecular dynamics simulation approach. J Clin Transl Res 2022;8:127-146. [pii]jctres.08.202202.003

[6] Sharma S, Deep S. In-silico drug repurposing for targeting SARS-CoV-2 main protease (M(pro)). J Biomol Struct Dyn 2022;40:3003–3010. https://doi.org/10.1080/07391102.2020.1844058

[7] Zhang D, Hamdoun S, Chen R et al. Identification of natural compounds as SARS-CoV-2 entry inhibitors by molecular docking-based virtual screening with bio-layer interferometry. Pharmacol Res 2021;172:105820. https://doi.org/10.1016/j.phrs.2021.105820

[8] Jang M, Park YI, Cha YE et al. Tea Polyphenols EGCG and Theaflavin Inhibit the Activity of SARS-CoV-2 3CL-Protease In Vitro. Evid Based Complement Alternat Med 2020;2020:5630838. https://doi.org/10.1155/2020/5630838

[9] Jang M, Park R, Park YI et al. EGCG, a green tea polyphenol, inhibits human coronavirus replication in vitro. Biochem Biophys Res Commun 2021;547:23–28. https://doi.org/10.1016/j.bbrc.2021.02.016

[10] Chiou WC, Chen JC, Chen YT et al. The inhibitory effects of PGG and EGCG against the SARS-CoV-2 3C-like protease. Biochem Biophys Res Commun 2021. https://doi.org/10.1016/j.bbrc.2020.12.106

[11] Park R, Jang M, Park YI et al. Epigallocatechin Gallate (EGCG), a Green Tea Polyphenol, Reduces Coronavirus Replication in a Mouse Model. Viruses 2021;13. https://doi.org/10.3390/v13122533

[12] Yang CC, Wu CJ, Chien CY, Chien CT. Green Tea Polyphenol Catechins Inhibit Coronavirus Replication and Potentiate the Adaptive Immunity and Autophagy-Dependent Protective Mechanism to Improve Acute Lung Injury in Mice. Antioxidants (Basel) 2021;10. https://doi.org/10.3390/antioX10060928 [doi]

[13] Hong S, Seo SH, Woo SJ, Kwon Y, Song M, Ha NC. Epigallocatechin Gallate Inhibits the Uridylate-Specific Endoribonuclease Nsp15 and Efficiently Neutralizes the SARS-CoV-2 Strain. J Agric Food Chem 2021;69:5948–5954. https://doi.org/10.1021/acs.jafc.1c02050

[14] Montone CM, Aita SE, Arnoldi A et al. Characterization of the Trans-Epithelial Transport of Green Tea (C. sinensis) Catechin Extracts with In Vitro Inhibitory Effect against the SARS-CoV-2 Papain-like Protease Activity. Molecules 2021;26.m https://doi.org/10.3390/molecules26216744

[15] Ngwe Tun MM, Luvai E, Nwe KM et al. Anti-SARS-CoV-2 activity of various PET-bottled Japanese green teas and tea compounds in vitro. Arch Virol 2022;167:1547–1557. https://doi.org/10.1007/s00705-022-05483-x

[16] Zhu Y, Xie DY. Docking Characterization and in vitro Inhibitory Activity of Flavan-3-ols and Dimeric Proanthocyanidins Against the Main Protease Activity of SARS-Cov-2. Front Plant Sci 2020;11:601316. https://doi.org/10.3389/fpls.2020.601316

[17] Kato Y, Higashiyama A, Takaoka E, Nishikawa M, Ikushiro S. Food phytochemicals, epigallocatechin gallate and myricetin, covalently bind to the active site of the coronavirus main protease in vitro. Adv Redox Res 2021;3:100021. https://doi.org/10.1016/j.arres.2021.100021

[18] Park J, Park R, Jang M, Park YI. Therapeutic Potential of EGCG, a Green Tea Polyphenol, for Treatment of Coronavirus Diseases. Life (Basel) 2021;11. https://doi.org/10.3390/life11030197

[19] Park J, Park R, Jang M, Park YI, Park Y. Coronavirus enzyme inhibitors-experimentally proven natural compounds from plants. J Microbiol 2022;60:347–354. https://doi.org/10.1007/s12275-022-1499-z

[20] Bimonte S, Forte CA, Cuomo M, Esposito G, Cascella M, Cuomo A. An Overview on the Potential Roles of EGCG in the Treatment of COVID-19 Infection. Drug Des Devel Ther 2021;15:4447–4454.https://doi.org/10.2147/DDDT.S314666

[21] Liu J, Bodnar BH, Meng F et al. Epigallocatechin gallate from green tea effectively blocks infection of SARS-CoV-2 and new variants by inhibiting spike binding to ACE2 receptor. Cell Biosci 2021;11:168.https://doi.org/10.1186/s13578-021-00680-8

[22] Kicker E, Tittel G, Schaller T, Pferschy-Wenzig EM, Zatloukal K, Bauer R. SARS-CoV-2 neutralizing activity of polyphenols in a special green tea extract preparation. Phytomedicine 2022;98:153970.https://doi.org/10.1016/j.phymed.2022.153970

[23] Momose Y, Maeda-Yamamoto M, Nabetani H. Systematic review of green tea epigallocatechin gallate in reducing low-density lipoprotein cholesterol levels of humans. Int J Food Sci Nutr 2016;67:606–613. https://doi.org/10.1080/09637486.2016.1196655

[24] Lin Y, Shi D, Su B et al. The effect of green tea supplementation on obesity: A systematic review and dose-response meta-analysis of randomized controlled trials. Phytother Res 2020. https://doi.org/10.1002/ptr.6697

[25] Kagawa Y. Influence of Nutritional Intakes in Japan and the United States on COVID-19 Infection. Nutrients 2022;14. https://doi.org/10.3390/nu14030633

[26] Khan N, Mukhtar H. Tea and health: studies in humans. Curr Pharm Des 2013;19:6141–6147. https://doi.org/10.2174/1381612811319340008

[27] Menegazzi M, Campagnari R, Bertoldi M, Crupi R, Di PR, Cuzzocrea S. Protective Effect of Epigallocatechin-3-Gallate (EGCG) in Diseases with Uncontrolled Immune Activation: Could Such a Scenario Be Helpful to Counteract COVID-19? Int J Mol Sci 2020;21. https://doi.org/10.3390/ijms21145171

[28] Abe SK, Inoue M. Green tea and cancer and cardiometabolic diseases: a review of the current epidemiological evidence. Eur J Clin Nutr 2020. https://doi.org/10.1038/s41430-020-00710-7

[29] Wessels I, Rolles B, Rink L. The Potential Impact of Zinc Supplementation on COVID-19 Pathogenesis. Front Immunol 2020;11:1712. https://doi.org/10.3389/fimmu.2020.01712

[30] Nanri A, Yamamoto S, Konishi M, Ohmagari N, Mizoue T. Green tea consumption and SARS-CoV-2 infection among staff of a referral hospital in Japan. Clin Nutr Open Sci 2022;42:1–5. https://doi.org/10.1016/j.nutos.2022.01.002

[31] Storozhuk MV. COVID -19: could green tea catechins reduce the risks? medRxiv 2020. https://doi.org/10.1101/2020.10.23.20218479

[32] Storozhuk MV. Green tea catechins against COVID-19? Lower COVID-19 morbidity and mortality in countries with higher per capita green tea consumption. Coronaviruses 2022. https://doi.org/10.2174/2666796703666220124103039

[33] Worldometers info. Coronavirus update. Available at: https://www.worldometersinfo/coronavirus/.

[34] Urashima M, Otani K, Hasegawa Y, Akutsu T. BCG Vaccination and Mortality of COVID-19 across 173 Countries: An Ecological Study. Int J Environ Res Public Health 2020;17.https://doi.org/10.3390/ijerph17155589

[35] World Bank. Population density (people per sq. km of land area). https://dataworldbankorg/indicator/EN POP DNST 2019.

[36] worldbank.org. Population ages 65 and above (% of total population). https://dataworldbankorg/indicator/SP POP 65UP TO ZS 2019.

[37] World Bank. Urban population (% of total population). https://dataworldbankorg/indicator/SP URB TOTL IN ZS Accessed 2019.

[38] UN. United Nations Development Program. Human Development Reports (2019). http://hdrundporg/en/content/2019-human-development-index-ranking 2019.

[39] Escobar LE, Molina-Cruz A, Barillas-Mury C. BCG vaccine protection from severe coronavirus disease 2019 (COVID-19). Proc Natl Acad Sci U S A 2020;117:17720–17726.https://doi.org/10.1073/pnas.2008410117

[40] Moore S, Hill EM, Tildesley MJ, Dyson L, Keeling MJ. Vaccination and non-pharmaceutical interventions for COVID-19: a mathematical modelling study. Lancet Infect Dis 2021;21:793–802.https://doi.org/10.1016/S1473-3099(21)00143-2

[41] Hellwig MD, Maia A. A COVID-19 prophylaxis? Lower incidence associated with prophylactic administration of ivermectin. Int J Antimicrob Agents 2021;57:106248. https://doi.org/10.1016/j.ijantimicag.2020.106248

[42] Annweiler C, Hanotte B, Grandin de lC, Sabatier JM, Lafaie L, Celarier T. Vitamin D and survival in COVID-19 patients: A quasi-experimental study. J Steroid Biochem Mol Biol 2020;204:105771. https://doi.org/10.1016/j.jsbmb.2020.105771

[43] Paiva L, Lima E, Motta. M, Marcone M, Baptista J. Investigation of the Azorean Camellia sinensis Processing Conditions to Maximize the Theaflavin 3,3?-di-O-Gallate Content as a Potential Antiviral Compound. Antioxidants 2022;11. https://doi.org/10.3390/antiox11061066

[44] ClinicalTrials gov Identifier: NCT04446065 20020. https://clinicaltrials.gov/ct2/show/NCT04446065

